# An audit assessing data quality, viral suppression, and transition to dolutegravir among children and adolescents with HIV in care at eThekwini Municipality, South Africa

**DOI:** 10.64898/2026.08.18.26359107

**Authors:** Minenhle Hlabisa, Luvo Mtila, Noluthando Lushaba, Kwena Tlhaku, Moherndran Archary, Johan S. van der Molen, Sanele Mbeje, Thokozani Khubone, Ntombifuthi Luthuli, Sharana Mahomed, Nigel Garrett, Lara Lewis, Jienchi Dorward, Yukteshwar Sookrajh, Jennifer Anne Brown

## Abstract

**Background:** The 2023 South African antiretroviral therapy (ART) guidelines recommend dolutegravir-based ART for children and adolescents with HIV (CAWH) >4 weeks old, including transition to dolutegravir-based ART if previously taking another regimen.

**Objectives:** We audited uptake of dolutegravir-based ART, viral load (VL) testing, and viral suppression among CAWH in care in eThekwini, South Africa. We also aimed to assess and improve the quality of routinely collected ART and VL data in the national HIV electronic register (TIER.Net) in this population.

**Methods:** We used TIER.Net line lists to identify CAWH aged ≤19 years in care in 54 eThekwini Municipality clinics between February and July 2025. CAWH who had died, transferred out, or were lost to follow-up were excluded. We reviewed clinical files and TIER.Net records simultaneously to compare all recorded ART regimens and recent (last 12 months) VL results. High or missing VLs were flagged for medical review, and data discrepancies were corrected.

**Results:** Among 3838 eligible CAWH, we reviewed files of 3379 (88%). 1991 (59%) were female and 2812 (83%) were aged 10-19 years. All 3379 (100%) were receiving dolutegravir-based ART. 193 (6%) had no recent VL result. Of those who did, 303 (10%) had a last VL ≥1000 copies/mL. We identified 941 (28%), 438 (13%), and 199 (6%) TIER.Net data capture errors for ART regimens, ART regimen start/stop dates, or recent VLs, respectively.

**Conclusion:** This audit at 54 facilities showed complete transition to dolutegravir among reviewed files of CAWH in care, but highlighted gaps in viral suppression and documentation.

## INTRODUCTION

Paediatric human immunodeficiency virus (HIV) remains a serious global health challenge, with children in low- and middle-income countries (LMICs) experiencing the greatest share of the disease burden. In South Africa, approximately 160 000 children under the age of 15 years were living with HIV in 2023^1^. Of these, 87% knew their HIV status, 73% of those who knew their status were on ART, and 75% of those on ART were virally suppressed^1^.

Treatment outcomes among children and adolescents are poorer than those observed among adults^2,3^. A lack of child-friendly ART formulations has long been a major contributing factor; many formulations are designed primarily for adults, and paediatric equivalents have frequently been delayed, unavailable, or costly. Additionally, some paediatric antiretroviral medications have an unpleasant taste or are challenging to administer to young children, which can lead to poor adherence^4^.

Since 2018, dolutegravir (DTG)-based ART has been designated as the preferred first- and second-line ART regimen in the World Health Organization (WHO) guidelines^5^. DTG was incorporated into South Africa’s national guidelines for adults, adolescents, and children weighing ≥20 kg in October 2019^6^. The South African 2023 ART guidelines subsequently extended DTG access to children weighing less than 20 kg, following the introduction of dispersible formulations; in practice, national rollout of DTG for this group began in late 2023^7^. This rollout was supported by the ODYSSEY trial, which showed that DTG–based regimens are more effective for children and adolescents when used as both first- and second-line therapy^8,9^, and by higher tolerability of the new 10 mg dispersible DTG capsules and existing DTG 50 mg tablets compared with prior regimens^4^.

In addition to appropriate ART regimens, accurate data capture is vital for monitoring paediatric HIV care and treatment outcomes, including evaluating the effects of the DTG transition on viral suppression. South African public clinics capture HIV data in the Three Interlinked Electronic Registers (TIER.Net) database, which is designed to facilitate the efficient and accurate collection of core data elements and key indicators necessary for monitoring HIV and ART services^10^. High-quality TIER.Net data furthermore supports clinical decision-making, alignment with national programme targets, and identification of gaps in care. By contrast, insufficient data quality can lead to misinformed decisions and compromised care. Regular data audits and validation processes are thus necessary to maintain data integrity and improve paediatric health outcomes.

This study aims to assess this transition, viral load (VL) outcomes, and TIER.Net data quality among children and adolescents living with HIV in the eThekwini Municipality of KwaZulu-Natal, South Africa.

## RESEARCH METHODS AND DESIGN

### Setting, study design, and population

KwaZulu-Natal has the second-highest HIV prevalence of all South African provinces^11^. We conducted a clinic audit with retrospective medical charts review at 54 facilities in eThekwini Municipality in KwaZulu-Natal. We aimed to assess clinical care, with a focus on DGT roll-out and treatment outcomes among children and adolescents in care for HIV, and to assess and improve TIER.Net data quality in this population. We included all CAWH who, at the time of facility audit, were aged ≤19 years and in care at the facility. We excluded CAWH who were recorded as dead, lost to follow-up, or transferred out in TIER.Net.

### Procedures

At each facility, we used TIER.Net, the national electronic medical record system, to generate line lists of eligible CAWH. TIER.Net contains information on demographic and clinical variables of people in care of HIV or tuberculosis^10^. The following procedures were followed in all the facilities.

In preparation for the audit, facility sub-district information officers and data clerks printed TIER.Net line lists of CAWH at the facilities and pulled the participants’ files

During the audit, the medical doctors conducted a detailed review of the participants’ files. This involved the audit team cross-checking each participant’s patient file against TIER.Net records, including all ART regimens with their respective start and stop dates as well as all VL results from the last 12 months with corresponding test dates. After documenting identified discrepancies between the clinical chart and TIER.Net records, data capturers were notified and corrections were implemented in real time. Any concerns, including lack of recent VL testing or elevated VLs without clinical follow-up, were flagged for the visiting medical doctor based at the facility. These patients would be booked for appointments with the visiting medical doctors to assess the patient and develop a management plan. General comments on areas for improvement, including clinical and operational performance and challenges, were recorded for reach facility.

### Outcomes

We assessed outcomes related to clinical outcomes, quality of care, and data quality. Clinical and quality of care outcomes included the proportion of CAWH receiving DTG-based ART, the proportion who did not have a documented VL result in the last 12 months, and, among those with at least one VL result in the last 12 months, the proportion with viraemia ≥1,000 copies/mL at their most recent measurement. Data quality outcomes included the proportion of CAWH with discrepancies in ART regimens and ART regimen start or stop dates between patient files and TIER.Net, as well as the proportion with VL results documented in patient files within the last 12 months that were not captured in TIER.Net. Finally, we retrospectively categorised areas for improvement noted during facility audits.

### Data management and analysis

Data were captured with tally sheets at each facility. We use descriptive statistics to summarize clinic level characteristics of children and adolescents in care for HIV in eThekwini Municipality, as well as to describe all endpoints. Continuous variables are summarized with medians and interquartile ranges (IQRs), categorical variables with frequencies and proportions.

### Spatial mapping

We visualised the geographic distribution of the assessed VL completeness and result outcomes across clinics through geospatial mapping using QGIS software version 3.40.0^12^. Mapping was performed using shapefile (vector layer) of clinic records containing clinic GPS coordinates, the proportion of CAWH with viraemia, and those without a recent VL across 54 eThekwini Municipality clinics, and the local municipality and ward boundaries shapefile (base map) covering areas where audit was done^13,14^. Inverse distance weighting interpolation (IDW) was applied to generate maps of these crude proportions, operating on the assumption that spatial phenomena at proximate locations are more similar than those further apart^15–17^.This spatial interpolation method estimates the proportions at unsampled locations (i.e. areas surrounding the audited clinics) by assigning weights that are inversely proportional to the power of the distance between known clinic proportions and estimated location. To preserve local spatial influence, we restricted the interpolation to a maximum of three nearest neighbouring clinics and used a default distance coefficient (a power parameter of 2).

### Ethical approval

The Strengthening Health systems through Audit and Programmatic data Evaluation (SHAPE) study within which this work was conducted was approved by the Biomedical Research Ethics Committee of the University of KwaZulu-Natal (BE646/17), the KwaZulu-Natal Provincial Health Research Ethics Committee (KZ_201807_021), and the eThekwini Municipality Health Department Research Committee, with a waiver of consent for analysis of de-identified, routinely collected data. This research is conducted in alignment with the Declaration of Helsinki.

## RESULTS

### Participant and clinic characteristics

Between 26 February 2025 and 17 July 2025, the audit was conducted at 54 eThekwini Municipality primary healthcare clinics. Clinic-level characteristics are shown in **Table 1**. Across the 54 clinics audited, the median (IQR) number of files audited was 58 (35-79), the median clinic-level proportion of female CAWH was 59% (51%-63%). In terms of age distribution, those aged 15-19 years represented a largest group across the clinics, with a median proportion of 56% (48%-60%).

**Table 1:**
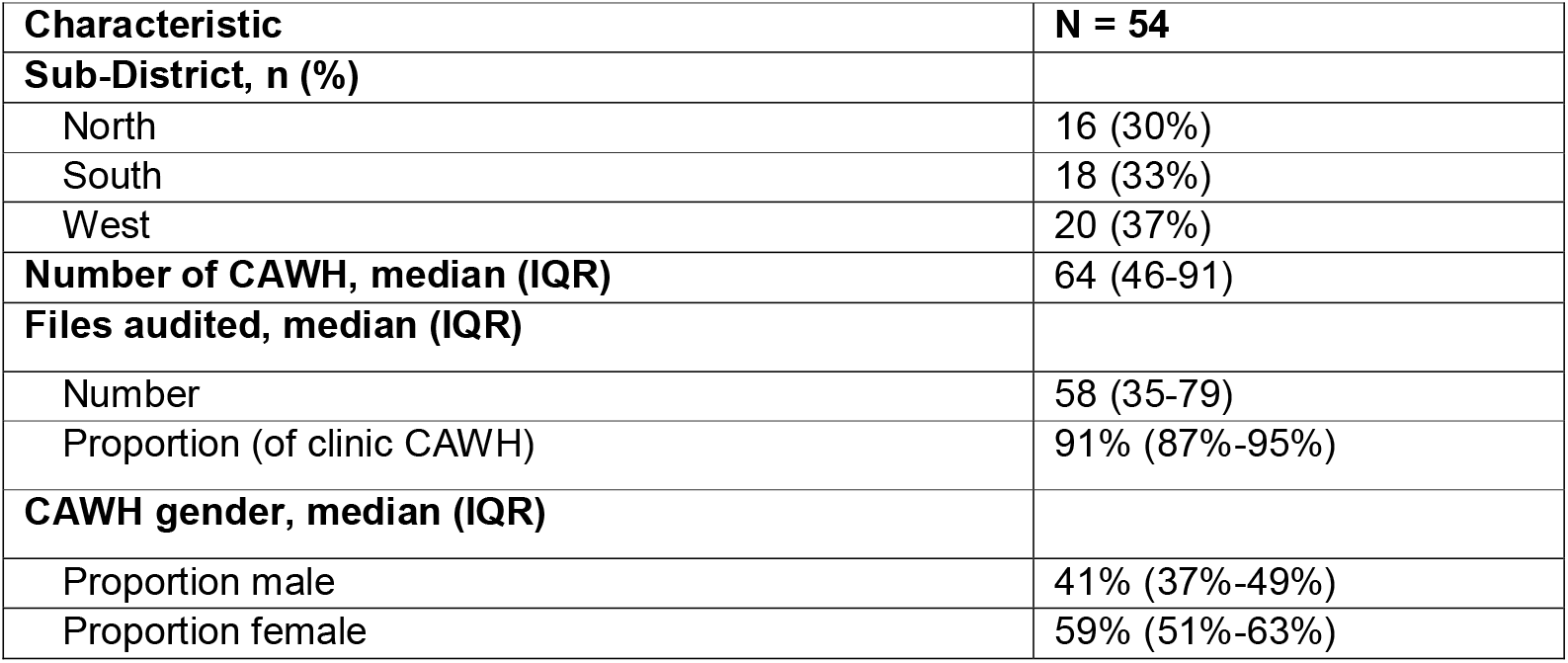
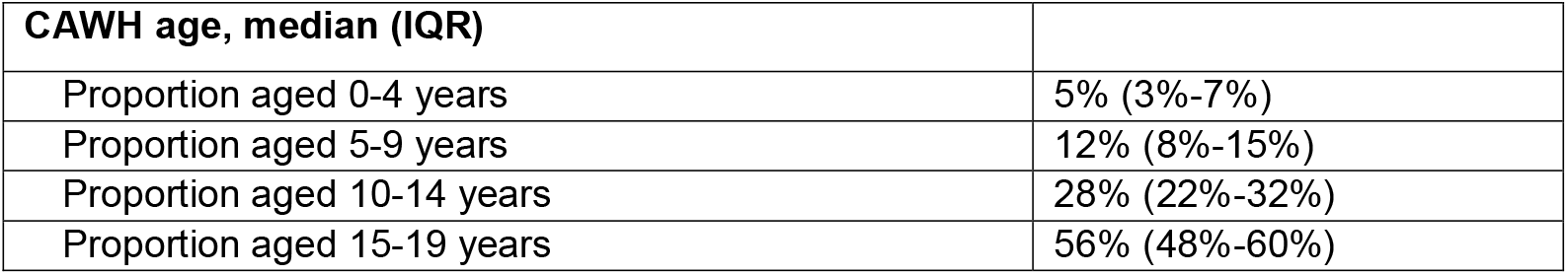
Characteristics of clinics audited between February and July 2025 in eThekwini Municipality. CAWH: children and adolescents with HIV; IQR: interquartile range.

3838 eligible CAWH were identified in TIER.Net, and files were reviewed for 3379 (88%). For the remaining 459 (12%) CAWH, files could not be found during the audit. Of CAWH whose files were reviewed, 1991 (59%) were female and 1388 (41%) were male. 162 (5%) were aged 0-4, 405 (12%) 5-9, 919 (27%) 10-14 and 1893 (56%) 15-19 years.

### Clinical, quality of care, and data quality outcomes

All 3379 (100%) participants were already receiving DTG. 193 (6%) did not have a recent VL result. Of those who did, 303 (10%) had a VL ≥1000 copies/mL at the latest measurement Additionally, low-level viraemia was recorded in 19 facilities in which 1169 files were audited; at these facilities, 60 (5%) did not have a recent VL and of the 1109 that did, 147 (13%) had a VL ≥1000 copies/mL and a further 123 (11%) had a VL of 50-999 copies/mL. Across all 54 facilities, the median clinic-level proportion with no VL in the last 12 months was 5% (IQR 2%-7%), and the median proportion with viraemia ≥1000 copies/mL among those with a recent VL result was 10% (6%-13%).

Spatial analysis of the audited clinics revealed a relatively homogeneous distribution in the proportion of CAWH without a recent VL test, with most areas demonstrating low to moderate monitoring gaps (Figure 1A). In contrast, the distribution of viraemia showed distinct spatial variation (Figure 1B). Higher levels of viraemia were observed in the far southern region of eThekwini, where the proportion was between 35% to 40%, and to a lesser degree in northern eThekwini.

**Figure 1.**
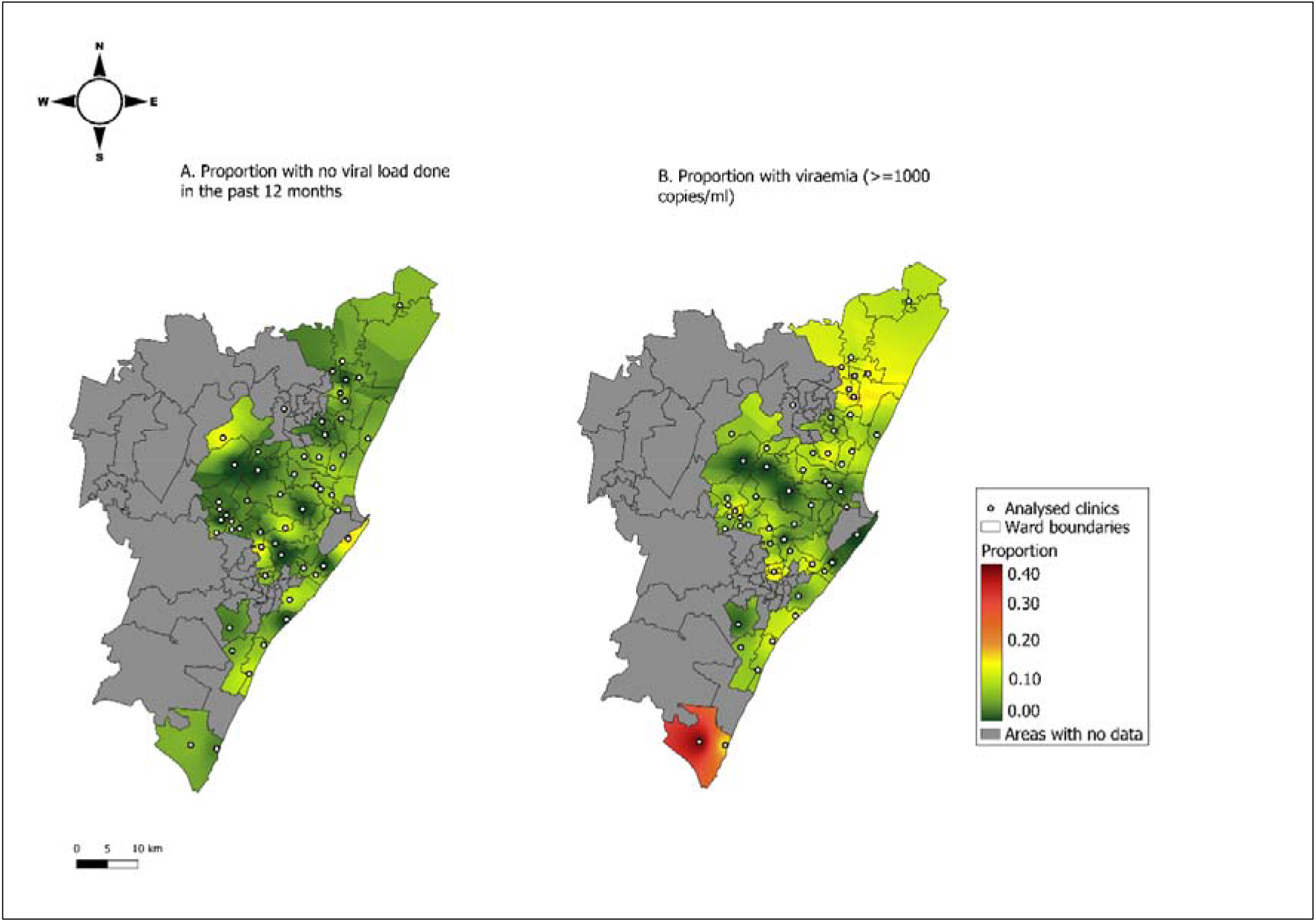
Spatial distribution of proportion of participants with no viral load in the past 12 months (left) and with viraemia ≥1000 copies/mL at the latest measurement in the past 12 months (right) in 54 facilities in eThekwini Municipality.

Overall, 941 (28%) had an ART regimen captured incorrectly in TIER.Net, 438 (13%) had ART regimen start/stop dates captured incorrectly, and 199 (6%) had VL results in the last 12 months that were not captured in TIER.Net. Across all facilities, the median clinic-level proportions with an incorrectly captured ART regimen, incorrectly captured ART start/stop dates, and recent VL data that was not captured in TIER.Net was 26% (13%-39%), 8% (4%-23%), and 4% (IQR 2%-8%), respectively.

### Further audit findings

Beyond the prespecified care and data quality endpoints, the most common areas flagged for improvement were low offering and/or poor recording of tuberculosis preventive treatment (flagged for 42 clinics), low provision and/or poor recording of family planning and of sexual and reproductive health services (34 clinics), inadequate referral of eligible CAWH to decentralised models of care for external ART pick-up as recommended by South African guidelines^7^ (28 clinics), and poor recording of anthropometric data (18 clinics), notably weight. Further common areas of concern included delayed action in case of viraemia (including low-level viraemia; 14 clinics), inadequate or disorganised documentation or duplicate files (13 clinics), and inadequate flagging of abnormal results (11 clinics; **Table 2**).

**Table 2:** Summary of identified gaps in HIV and tuberculosis care across audited facilities.

| **Gaps in care** | **Number of clinics affected** |
| --- | --- |
| **Prevention and screening** | **44** |
| Inadequate tuberculosis preventive therapy offering or capturing | 42 |
| Inadequate offering of sexual and reproductive health / family planning | 34 |
| Inadequate syphilis screening | 2 |
| High number of teenage pregnancies | 1 |
| **Decentralised ART pick-up** | **31** |
| Inadequate referral of eligible CAWH | 28 |
| Referral despite ineligibility | 5 |
| **Clinical monitoring and management** | **27** |
| Poor documentation of anthropometric data (notably weight) | 18 |
| Delayed action in case of viraemia | 14 |
| Lack of tracing | 2 |
| Inadequate enhanced adherence counselling | 1 |
| **Laboratory monitoring and results management** | **15** |
| Poor flagging of abnormal results | 11 |
| CD4% not recorded for children <5 years | 2 |
| Poor baseline CD4 capturing | 1 |
| CD4 monitoring without indication | 1 |
| **ART treatment and documentation** | **11** |
| Late switch to TDF/3TC/DTG upon meeting weight and age criteria | 7 |
| Incorrect capturing of first- and second-line ART | 4 |
| DTG boosting in case of tuberculosis treatment not documented | 1 |
| ART transition without clinical assessment | 1 |
| **Documentation and file management** | **16** |
| General poor / disorganised documentation | 8 |
| Duplicate files | 5 |
| Next visit dates unclear | 5 |
| Incorrect date of birth | 1 |

3TC: lamivudine; ART: antiretroviral therapy; CAWH: children and adolescents with HIV; DTG: dolutegravir; TDF: tenofovir disoproxil fumarate.

## DISCUSSION

In this retrospective audit of medical charts from 54 facilities in eThekwini Municipality conducted from February to July 2025, we found that the paediatric transition to DTG was successfully and universally implemented. Most participants (94%) had a recent VL test, and of these, 90% had an HIV VL <1000 copies/mL. However, the audit also showed discrepancies between clinical files and the TIER.Net electronic HIV register, particularly regarding ART regimens. Furthermore, it revealed gaps in service delivery, notably in the provision of tuberculosis preventive treatment and sexual and reproductive health services, as well as in broader documentation.

The 10% proportion of CAWH in active care with viraemia ≥1000 copies/mL in our audit is lower than recently reported for children and adolescents <15 years in South Africa by UNAIDS^1^ or in a recent systematic review among CAWH receiving ART in Africa^18^. However, it still falls short of the UNAIDS 95% viral suppression target. With regards to data quality, a previous analysis of TIER.Net data quality involving record review and participant tracing identified overreporting of loss to follow-up and underreporting of death and clinic transfers in TIER.Net data^19^. More recently, our group compared TIER.Net data with national databases for laboratory results and ART prescription. Findings suggested high alignment of clinic-level data on referral to decentralized ART programmes and VL results, but underreporting of CD4 count capture in TIER.Net^20^. In the present audit, in addition to discrepancies between patient files and TIER.Net, we report gaps care and documentation in patient files including duplicate files, poor recording of anthropometric measurements, and inadequate flagging of abnormal results. These findings reflect persistent weaknesses in routine clinical documentation and health information management systems.

This audit had several limitations. First, we assessed participant files only for individuals retained in care. Second, low-level viraemia was not systematically captured across all facilities. Third, factors such as medication adherence and socioeconomic status were not measured. Fourth, there was no direct contact with participants, which limited the ability to verify discrepancies in the records. Finally, the IDW approach used for geospatial mapping relies strictly on the distance and is susceptible to the “bull’s eye” effect, where isolated clinic data points circular artifacts that may not reflect true continuous geographic transition.

Despite these limitations, the audit provided critical feedback on the CAWH programme and had direct operational benefits. Patients with high or missing VLs were flagged for timely clinical follow-up, and discrepancies in ART regimens and treatment dates were corrected in TIER.Net, improving the accuracy of programme data. The process also engaged clinic staff in reviewing documentation practices, reinforcing the importance of complete and accurate data capture.

## CONCLUSION

Routine audits can strengthen both clinical care and health information systems in high-burden HIV settings. This audit demonstrates a successful transition of CAWH to DTG-based ART in eThekwini in alignment with national treatment guidelines. However, gaps in viral suppression, VL monitoring, as well as inaccuracies in routine electronic and paper-based data capture persist and limit optimal clinical management. This needs to be addressed through strengthened VL follow-up and ongoing support for accurate TIER.Net documentation to sustain treatment gains and improve health outcomes for CAWH.

## Author contributions

JD and YS conceptualised the study. YS, LM, NL, KT, and MH oversaw data collection. YS, LM, NL, KT, MA, JSvdM, SaM, TK, ShM, NG, LL, JD and JAB were responsible for various components of project administration. MH, SaM and JAB analysed the data. MH and JAB drafted the manuscript. MH, YS, JAB and JD had full access to all data in the study and final responsibility to submit for publication. All authors contributed to interpretation of results, critically reviewed and edited the manuscript, and consented to final publication.

## Acknowledgements

We thank the eThekwini Municipality as well as the staff and clients in care at all participating healthcare facilities.

## Data sharing

Data are available from the corresponding author upon reasonable request.

